# Incubation Period and Other Epidemiological Characteristics of 2019 Novel Coronavirus Infections with Right Truncation: A Statistical Analysis of Publicly Available Case Data

**DOI:** 10.1101/2020.01.26.20018754

**Authors:** Natalie M. Linton, Tetsuro Kobayashi, Yichi Yang, Katsuma Hayashi, Andrei R. Akhmetzhanov, Sung-mok Jung, Baoyin Yuan, Ryo Kinoshita, Hiroshi Nishiura

**Affiliations:** Graduate School of Medicine, Hokkaido University, Sapporo, Hokkaido, Japan; CREST, Japan Science and Technology Agency, Honcho 4-1-8, Kawaguchi, Saitama 332-0012, Japan

**Keywords:** epidemiology, incubation period, virus, distribution, emerging infectious diseases

## Abstract

The geographic spread of 2019 novel coronavirus (COVID-19) infections from the epicenter of Wuhan, China, has provided an opportunity to study the natural history of the recently emerged virus. Using publicly available event-date data from the ongoing epidemic, the present study investigated the incubation period and other time intervals that govern the epidemiological dynamics of COVID-19 infections. Our results show that the incubation period falls within the range of 2–14 days with 95% confidence and has a mean of around 5 days when approximated using the best-fit lognormal distribution. The mean time from illness onset to hospital admission (for treatment and/or isolation) was estimated at 3–4 days without truncation and at 5–9 days when right truncated. Based on the 95th percentile estimate of the incubation period, we recommend that the length of quarantine should be at least 14 days. The median time delay of 13 days from illness onset to death (17 days with right truncation) should be considered when estimating the COVID-19 case fatality risk.

## 1 Introduction

As of 31 January 2020, mainland China reported 11,791 confirmed cases of novel coronavirus (COVID-19) infections, causing 259 deaths [1]. Initially, these infections were thought to result from zoonotic (animal-to-human) transmission; however, recently published evidence [2] and the exponential growth of case incidences show compelling evidence of human-to-human secondary transmission fueled by travel, with many cases detected in other parts of the world [3]. This geographic expansion beyond the initial epicenter of Wuhan provides an opportunity to study the natural history of COVID-19 infection, as these migration events limit the risk of infection to the time during which an individual traveled to an area where exposure could occur [4].

The incubation period is defined as the time from infection to illness onset. Knowledge of the incubation period of a directly transmitted infectious disease is critical to determine the time period required for monitoring and restricting the movement of healthy individuals (i.e., the quarantine period) [5, 6]. The incubation period also aids in understanding the relative infectiousness of COVID-19 and can be used to estimate the epidemic size [7].

Time-delay distributions including the dates of hospital admission (for treatment and/or isolation) and death also inform the temporal dynamics of epidemics. A published clinical study on the COVID-19 epidemic has already shown that the average time delay from illness onset to hospital admission is approximately 7 days [8], but this distribution has yet to be explicitly estimated. The time from hospital admission to death is also critical to the avoidance of underestimation when calculating case fatality risk [9]. Using publicly available data from the ongoing epidemic of COVID-19 with known event dates, the present study aimed to estimate the incubation period and other time intervals that govern the interpretation of epidemiological dynamics of COVID-19 infections.

## 2 Methods

### 2.1 Epidemiological data

We retrieved information on cases with confirmed COVID-19 infection and diagnosis outside of the epicenter of Wuhan, China, based on official reports from governmental institutes, as well as reports on deceased cases from both in and outside of Wuhan. We aggregated the data directly from government websites or from news sites that quoted government statements. The data were collected in real time, and thus may have been updated as more details on cases became available. The arranged data include a selection of cases reported through 31 January 2020 and are available as Supplementary Tables S1 and S2.

Specifically, we collected the dates of exposure (entry and/or exit from Wuhan or dates of close contact with a Wuhan resident/known epidemic case), illness onset, earliest healthcare seeking related to infection, hospital admission (for treatment and/or isolation), and death. Cases included both residents from other locations who travelled to Wuhan, as well as individuals who lived, worked, or studied in Wuhan (hereafter: Wuhan residents) but who were diagnosed outside of Wuhan and reported by the governments of the locations where their infection was detected. We thus estimated the incubation period by (i) excluding Wuhan residents and (ii) including Wuhan residents. The former may be more precise in defining the interval of exposure, but the sample size is greater for the latter. More detailed information about the criteria used for the estimation of each defined time interval and data used are described in Supplementary Text S1.

### 2.2 Statistical model

We used the dates of three critical points in the course of infection–symptom onset, hospital admission, and death–to calculate four time intervals: the time from (a) exposure to illness onset (i.e., incubation period), (b) illness onset to hospital admission, (c) illness onset to death, and (d) hospital admission to death. We used a doubly interval-censored [10] likelihood function to estimate the parameter values for these intervals, written as:

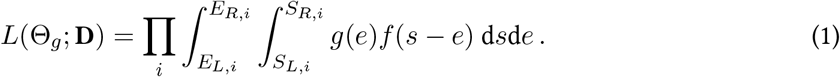

Here, in the case of (a) *g*(.) is the probability density function (PDF) of exposure following a uniform distribution, and *f* (.) is the PDF of the incubation period independent of *g*(.). **D** represents a dataset among all observed cases *i*, where exposure and symptom onset fall within the lower and upper bounds (*E*_*L*_, *E*_*R*_) and (*S*_*L*_, *S*_*R*_). We fit the PDF *f* (.) to lognormal, Weibull, and gamma distributions.

To address the selection bias in the dataset due to the continued growth of the outbreak (i.e., cases with shorter incubation periods are more likely to be included in the dataset), we also accounted for right truncation using the formula:

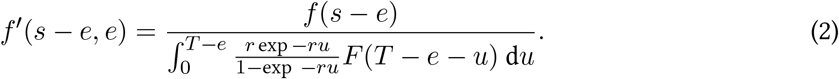

Here, *r* is the exponential growth rate (estimated at 0.14 [11]), and *T* is the latest time of observation (31 January 2020), and *F* (.) is the cumulative density function of *f* (.).

In both cases, we used Bayesian methods to infer parameter estimates and obtain credible intervals. We selected the best fit model by using the widely applicable information criterion (WAIC). We also verified that the Bayesian estimates were in line with pointwise estimates derived by maximum likelihood estimation (MLE). As the formulation of the likelihood with right truncation (1)–(2) contained the function *f* ^*′*^ and was dependent on both the time interval (*s − e*) and time of exposure *e*, we generalized a previously obtained result for doubly interval-censored likelihood with *f* ^*′*^(*s − e, e*) ≡ *f* (*s − e*) [10].

The data were processed using R version 3.6.2 [12], MLE was computed using Julia version 1.3 [13], and the Markov chain Monte Carlo (MCMC) simulations were performed in Stan (cmdStan version 2.22.1 [14]). All code is freely available at the github repository [15].

## 3 Results

The ratio of male to female cases among living cases resembled [2], at 58%, with most 30–59 years of age (information missing for 9 cases). The deceased cases were more predominantly male (70%) and older (85% were 60 years of age or older). Table 1 shows estimates for the various time intervals without right truncation. For the incubation period estimates, the lognormal distribution provided the best fit to the data, both when excluding and including Wuhan residents. The mean incubation period was estimated at 5.0 days (95% credible interval [CI]: 4.2, 6.0) when excluding Wuhan residents (n = 52) and 5.6 days (95% CI: 5.0, 6.3) when including Wuhan residents (n = 158).

**Table 1.** Incubation period and other time-delay distributions for COVID-19 outbreak cases reported in January 2020. WR: Wuhan residents. SD: standard deviation. WAIC: widely applicable information criterion. Weights can be interpreted as the probability of each model given the data and were calculated using stacking method [21].

|  | Incubation Period<br>Excluding WR<br>(Days) | Incubation Period<br>Including WR<br>(Days) | Onset to Hospital<br>Admission, Living<br>(Days) | Onset to Hospital<br>Admission,<br>Deceased (Days) | Onset to Death<br>(Days) | Hospital<br>Admission to<br>Death (Days) |
| --- | --- | --- | --- | --- | --- | --- |
| Number of cases | 52 cases | 158 cases | 155 cases | 34 cases | 34 cases | 39 cases |
| Lognormal |  |  |  |  |  |  |
| Mean | 5.0 (4.2, 6.0) | 5.6 (5.0, 6.3) | 3.9 (2.9, 5.3) | 6.2 (5.0, 7.8) | 14.5 (12.5, 17.0) | 8.6 (6.8, 10.8) |
| SD | 3.0 (2.1, 4.5) | 2.8 (2.2, 3.6) | 9.1 (5.2, 16.3) | 4.3 (2.9, 6.6) | 6.7 (4.9, 9.4) | 6.7 (4.5, 10.3) |
| 5% | 1.7 (1.2, 2.3) | 2.3 (1.9, 2.7) | 0.2 (0.1, 0.3) | 1.9 (1.2, 2.5) | 6.5 (4.9, 7.9) | 2.2 (1.5, 3.0) |
| Median | 4.3 (3.5, 5.1) | 5.0 (4.4, 5.6) | 1.5 (1.2, 1.9) | 5.1 (4.1, 6.3) | 13.2 (11.3, 15.3) | 6.7 (5.3, 8.3) |
| 95% | 10.6 (8.5, 14.1) | 10.8 (9.3, 12.9) | 14.0 (10.3, 20.1) | 13.9 (10.8, 19.6) | 26.8 (22.3, 34.5) | 20.5 (15.7, 28.7) |
| 99% | 15.4 (11.7, 22.5) | 14.9 (12.3, 18.7) | 35.0 (23.2, 56.9) | 21.1 (15.3, 32.8) | 36.0 (28.6, 49.8) | 32.6 (23.3, 50.1) |
| WAIC | 257.2 | 859.6 | 693.5 | 183.9 | 221.9 | 240.1 |
| Weight | 1.00 | 1.00 | 0.00 | 0.33 | 1.00 | 0.02 |
| Weibull |  |  |  |  |  |  |
| Mean | 5.4 (4.3, 6.6) | 5.8 (5.2, 6.5) | 3.4 (2.7, 4.2) | 6.5 (5.2, 8.0) | 15.1 (12.7, 17.8) | 8.9 (7.3, 10.4) |
| SD | 3.6 (2.8, 4.7) | 2.8 (2.3, 3.5) | 4.4 (3.3, 6.0) | 4.0 (3.1, 5.4) | 7.6 (6.1, 9.7) | 5.4 (4.2, 7.3) |
| 5% | 0.9 (0.5, 1.4) | 2.1 (1.5, 2.6) | 0.1 (0.0, 0.1) | 1.9 (1.2, 2.5) | 4.2 (2.5, 6.0) | 1.7 (0.9, 2.7) |
| Median | 4.7 (3.6, 5.8) | 5.3 (4.7, 6.0) | 1.8 (1.4, 2.3) | 5.1 (4.1, 6.3) | 14.3 (11.8, 17.1) | 8.0 (6.2, 9.8) |
| 95% | 12.0 (9.8, 15.6) | 11.0 (9.6, 12.9) | 11.7 (9.3, 15.6) | 13.9 (10.8, 19.6) | 28.6 (24.5, 34.9) | 18.8 (15.5, 24.4) |
| 99% | 15.9 (12.8, 21.5) | 14.2 (12.1, 17.0) | 20.3 (15.3, 29.2) | 21.1 (15.3, 32.8) | 35.0 (29.5, 44.4) | 24.2 (19.5, 33.0) |
| WAIC | 273.8 | 871.8 | 662.4 | 185.5 | 231.3 | 236.1 |
| Weight | 0.00 | 0.00 | 0.00 | 0.29 | 0.00 | 0.96 |
| Gamma |  |  |  |  |  |  |
| Mean | 5.3 (4.3, 6.6) | 6.0 (5.3, 6.7) | 3.3 (2.7, 4.0) | 6.5 (5.2, 8.0) | 15.0 (12.8, 17.5) | 8.8 (7.2, 10.8) |
| SD | 3.2 (2.4, 4.3) | 3.1 (2.7, 3.7) | 4.2 (3.3, 5.4) | 4.0 (2.9, 5.6) | 6.9 (5.2, 9.1) | 5.7 (4.3, 7.8) |
| 5% | 1.4 (0.7, 2.0) | 1.5 (1.1, 2.0) | 0.0 (0.0, 0.1) | 1.6 (0.8, 2.4) | 5.8 (3.9, 7.6) | 1.9 (1.0, 2.9) |
| Median | 4.7 (3.8, 5.7) | 5.6 (4.9, 6.4) | 1.7 (1.2, 2.2) | 5.6 (4.5, 6.9) | 13.9 (11.8, 17.1) | 7.6 (6.0, 9.3) |
| 95% | 11.3 (9.2, 14.5) | 11.7 (10.3, 13.4) | 11.6 (9.4, 14.7) | 13.8 (11.1, 18.6) | 27.4 (23.2, 34.1) | 19.9 (15.7, 25.7) |
| 99% | 15.2 (12.0, 20.2) | 14.5 (12.6, 16.9) | 19.4 (15.4, 25.1) | 18.6 (14.6, 26.2) | 34.9 (28.8, 44.8) | 26.8 (21.1, 36.5) |
| WAIC | 265.8 | 895.9 | 656.6 | 183.4 | 225.1 | 236.3 |
| Weight | 0.00 | 0.00 | 1.00 | 0.38 | 0.00 | 0.02 |

The median time from illness onset to hospital admission was estimated at 3.3 days (95% CI: 2.7, 4.0) among living cases and 6.5 days (95% CI: 5.2, 8.0) among deceased cases using the gamma distribution, which provided the best fit for both sets of data. Figure 1A shows the corresponding PDFs. Data from the time from illness onset and hospital admission to death best fit lognormal and Weibull distributions, respectively, as presented in Figure 1B,C. The mean time from illness onset to death was 15.0 days (95% CI: 12.8, 17.5) and from hospital admission to death was 8.8 days (95% CI: 7.2, 10.8).

**Figure 1.**
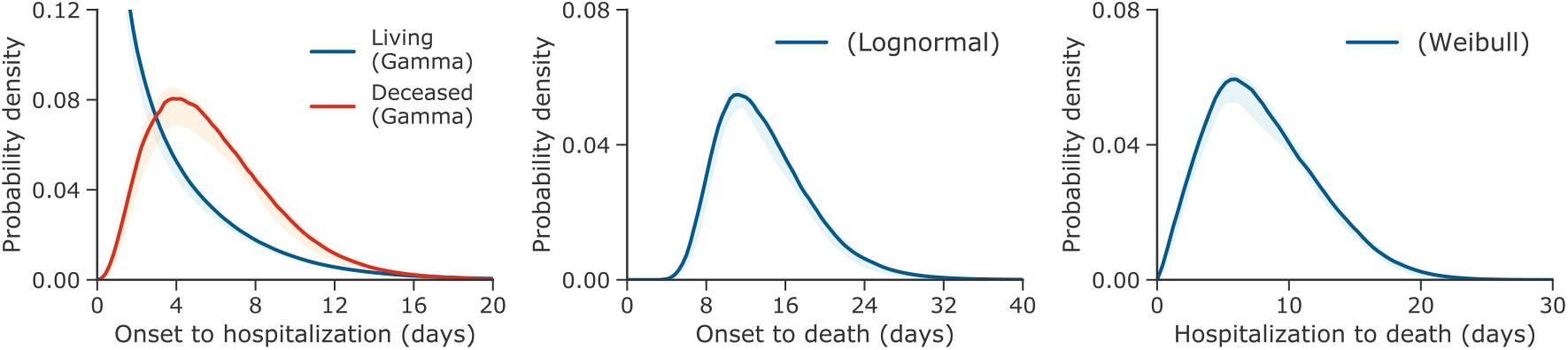
Probability distributions of the time from illness onset or hospital admission to hospital admission or death for COVID-19 outbreak cases reported through 31 January 2020. (**A**) Probability density of the time from illness onset to hospital admission in days set to the best-fit gamma distribution. (**B**) Probability density of the time from illness onset to death in days set to the best-fit lognormal distribution. (**C**) Probability density of the time from hospital admission to death in days set to the best-fit Weibull distribution.

Table 2 shows estimates for the fit of the lognormal distribution for each interval when accounting for right truncation. The mean incubation period was 5.6 days (95% CI: 4.4, 7.4) when excluding Wuhan residents–slightly larger than the estimate without right truncation. The mean estimate for illness onset to hospital admission was 9.7 days (95% CI: 5.4, 17.0) for living cases and 6.6 days (95% CI: 5.2, 8.8) for deceased cases, with the former nearly 2.5 times the length of its untruncated version. Illness onset to death and hospital admission to death were likewise longer than their non-truncated counterparts, at 20.2 days (95% CI: 15.1, 29.5) and 13.0 days (95% CI: 8.7, 20.9), respectively.

**Table 2.**
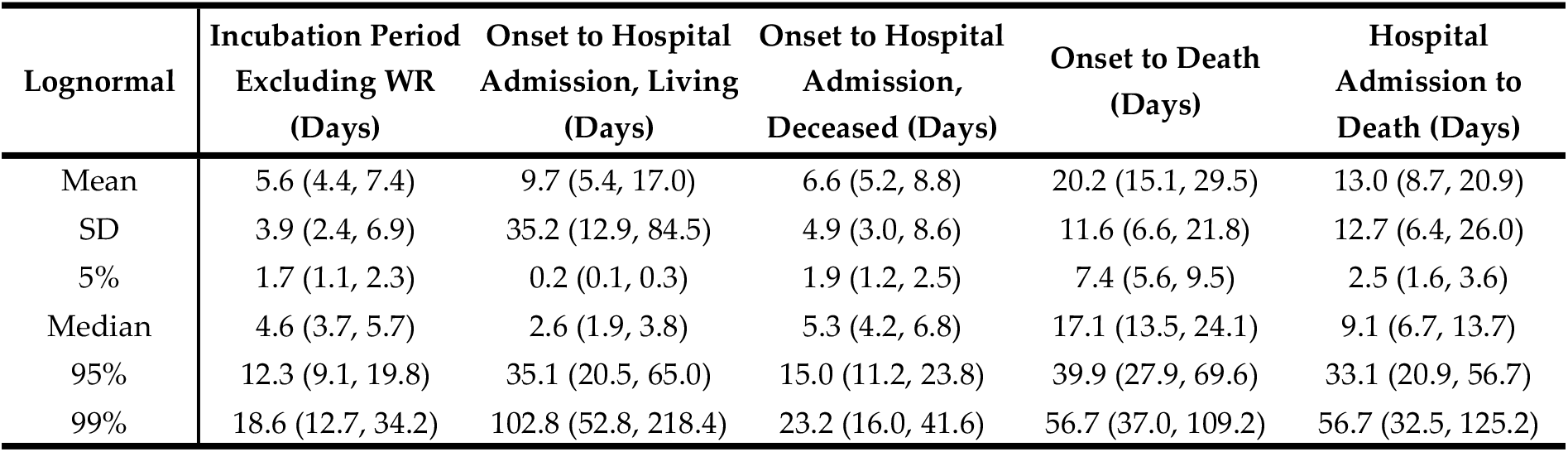
Right-truncated incubation period and other time-delay distributions for COVID-19 outbreak cases reported in January 2020 applied to the lognormal distribution. WR: Wuhan residents. SD: standard deviation.

Figure 2 shows the cumulative distribution function of the incubation period with and without right truncation. The 5th and 95th percentiles are shown in addition to the median. The 95th percentiles were estimated at 10.6 days (95% CI: 8.5, 14.1) and 10.8 days (95% CI: 9.3, 12.9) for non-truncated data excluding and including Wuhan residents and 12.3 days (95% CI: 9.1, 19.8) when applying right truncation and excluding Wuhan residents. The respective median values for these CDFs were 4.3 days (95% CI: 3.5, 5.1), 5.0 days (95% CI: 4.4, 5.6), and 4.6 days (95% CI: 3.7, 5.7).

**Figure 2.**
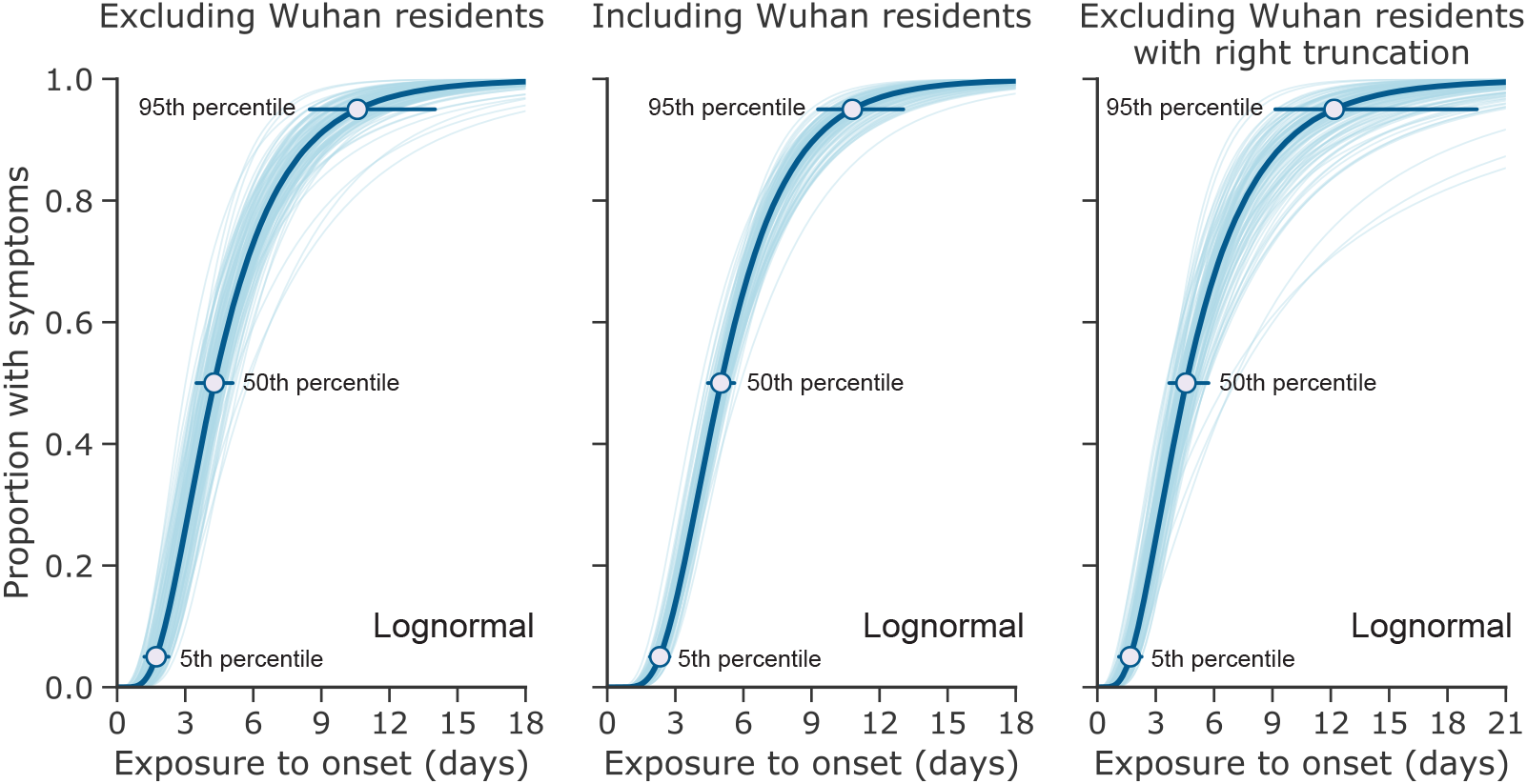
Estimated cumulative distribution for the incubation period of COVID-19 infections from outbreak cases reported through 31 January 2020. The data are from public case reports. Left and center: non-truncated estimates excluding (*n* = 52) and including (*n* = 158) Wuhan residents. Right: right-truncated estimates excluding Wuhan residents (*n* = 52).

## 4 Discussion

The present study advances the public discussion on COVID-19 infections by presenting explicit estimations of the incubation period and other epidemiologic characteristics using publicly available data. Our estimated mean incubation period of approximately 5 days is comparable to known mean values of the incubation period for severe acute respiratory syndrome (SARS) and Middle East respiratory syndrome (MERS) [9, 16–18], as well as other recent estimates of the incubation period for COVID-19 [18]. In addition to empirically showing the comparability of COVID-19 to other disease-causing coronaviruses, the present study has also shown that the 95th percentile of the incubation period is around 10–14 days, indicating that a 14-day quarantine period would largely ensure the absence of disease among healthy exposed individuals.

Wuhan residents have a less precisely defined exposure period compared to travelers and secondary cases from known human to human transmission events. However, our calculations have shown that adding more cases to the dataset even with uncertainty reduces both the variance of the estimates and selection bias, improving the fit of the distribution mean. Our estimates are in agreement with the report of Li et al. [2]. A recent study by Backer et al. [18] noted a similar finding in their analysis of the incubation period for 88 cases (including 63 Wuhan residents). However, the estimates of Backer et al. for the model that included Wuhan residents were subject to overestimation as the lower bounds for Wuhan residents–who had unknown left exposure dates–were fixed in their analysis. In contrast, we considered the left exposure dates for Wuhan residents as parameters to be fitted—see [14] for details. Notably, our results demonstrated the overall benefit of using additional case data, even when some of exposure values were not precisely known.

The time from the illness onset to death is also comparable to SARS [16], and the 15–20-day mean delay indicates that a crude estimation of the ratio of the cumulative number of deaths to that of confirmed cases will tend to result in an underestimation of the case fatality risk, especially during the early stage of epidemic spread. During the SARS epidemic in Hong Kong, 2003, the time from illness onset to hospital admission was shown to have shortened as a function of the calendar time, gradually reflecting the effects of contact tracing [9]. It remains to be seen if this will be the case for COVID-19 as well. The time delay distribution between illness onset and hospital admission may also be negatively associated with the basic reproduction number, i.e., the average number of secondary cases generated by a single primary case in a fully susceptible population [19].

The median time from illness onset to hospital admission was approximately 4 days among cases not known to be deceased at the time of the case report, and 6 days among cases reported as deceased. The reasons for this difference are not altogether clear. However, the living cases include persons who were isolated–in some cases more for reducing transmission than for treatment purposes–while all deceased cases were admitted for treatment. In addition, deceased cases for whom information was available had onset dates closer to the beginning of the outbreak compared to the living cases, who mostly had onset in the latter two-thirds of January 2020. The time delay distributions from illness onset to hospital admission for cases reported later in the epidemic, when there was a more widespread recognition of the virus and a more prevalent social imperative for those with symptoms to seek healthcare, may differ from those of early cases [2].

Several limitations of the present study exist. First, the dataset relies on publicly available information that is not uniformly distributed (i.e., collected from various sources), and therefore the availability of dates relevant to our analyses is limited to a small, selective sample that is not necessarily generalizable to all confirmed cases. Moreover, given the novelty of the COVID-19 pneumonia, it is possible that illness onset and other event data were handled differently between jurisdictions (e.g., was illness onset the date of fever or date of dyspnea?). Second, our data include very coarse date intervals with some proxy dates used to determine the left and/or right hand dates of some intervals. Third, as the sample size was limited, the variance is likely to be biased. Fourth, we were not able to examine the heterogeneity of estimates by different attributes of cases (e.g., severity of disease) [19]. Lastly, as we only have information on confirmed cases, there is a bias towards more severe disease–particularly for earlier cases.

This study presents the estimates of epidemiological characteristics of COVID-19 infections that are key parameter for studies on incidence, case fatality, and epidemic final size, among other possibilities [7, 11]. From the 95th percentile estimate of the incubation period we found that the length of quarantine should be at least 14 days, and we stress that the 17–24-day time delay from illness onset to death must be addressed when estimating COVID-19 case fatality risk. This study was made possible only through open sharing of case data from China and other countries where cases were diagnosed. Continued communication of dates and other details related to exposure and infection is crucial to furthering scientific understanding of the virus, the infections it causes, and preventive measures that can be used to contain and mitigate epidemic spread.

## Data Availability

Used dataset is available as the Supplementary Material

## Supplementary material

Supplementary table 1: Event dates for exported cases included in the analysis, Supplementary table 2: Event dates for deceased cases included in the analysis, Supplementary Text 1, Estimation of the time interval distribution using doubly interval-censored likelihood, estimation of the time interval distributions using Bayesian framework, and data cleaning rules implemented for the various time intervals.

## Author Contributions

N.M.L., T.K., A.R.A., and H.N. conceived the study and participated in the study design. All authors assisted in collecting the data. N.M.L., T.K. and H.N. analyzed the data and T.K., H.N., N.M.L. and Y.Y. drafted the manuscript. All authors edited the manuscript and approved the final version.

